# Structural brain abnormalities and aggressive behaviour in schizophrenia: Mega-analysis of data from 2095 patients and 2861 healthy controls via the ENIGMA consortium

**DOI:** 10.1101/2024.02.04.24302268

**Authors:** Jelle Lamsma, Adrian Raine, Seyed M. Kia, Wiepke Cahn, Dominic Arold, Nerisa Banaj, Annarita Barone, Katharina Brosch, Rachel Brouwer, Arturo Brunetti, Vince D. Calhoun, Qian H. Chew, Sunah Choi, Young-Chul Chung, Mariateresa Ciccarelli, Derin Cobia, Sirio Cocozza, Udo Dannlowski, Paola Dazzan, Andrea de Bartolomeis, Marta Di Forti, Alexandre Dumais, Jesse T. Edmond, Stefan Ehrlich, Ulrika Evermann, Kira Flinkenflügel, Foivos Georgiadis, David C. Glahn, Janik Goltermann, Melissa J. Green, Dominik Grotegerd, Amalia Guerrero-Pedraza, Minji Ha, Elliot L. Hong, Hilleke Hulshoff Pol, Felice Iasevoli, Stefan Kaiser, Vasily Kaleda, Andriana Karuk, Minah Kim, Tilo Kircher, Matthias Kirschner, Peter Kochunov, Jun Soo Kwon, Irina Lebedeva, Rebekka Lencer, Tiago R. Marques, Susanne Meinert, Robin Murray, Igor Nenadić, Dana Nguyen, Godfrey Pearlson, Fabrizio Piras, Edith Pomarol-Clotet, Giuseppe Pontillo, Stéphane Potvin, Adrian Preda, Yann Quidé, Amanda Rodrigue, Kelly Rootes-Murdy, Raymond Salvador, Antonin Skoch, Kang Sim, Gianfranco Spalletta, Filip Spaniel, Frederike Stein, Florian Thomas-Odenthal, Andràs Tikàsz, David Tomecek, Alexander Tomyshev, Mario Tranfa, Uyanga Tsogt, Jessica A. Turner, Theo G. M. van Erp, Neeltje E. M. van Haren, Jim van Os, Daniela Vecchio, Lei Wang, Adrian Wroblewski, Thomas Nickl-Jockschat

**Author notes:** Corresponding author; Department of Criminology, Vrije Universiteit Amsterdam, De Boelelaan 1105, 1081 HV Amsterdam, the Netherlands.

## Abstract

**Background:** Schizophrenia is associated with an increased risk of aggressive behaviour, which may partly be explained by illness-related changes in brain structure. However, previous studies have been limited by group-level analyses, small and selective samples of inpatients and long time lags between exposure and outcome.

**Methods:** This cross-sectional study pooled data from 20 sites participating in the international ENIGMA-Schizophrenia Working Group. Sites acquired T1-weighted and diffusion-weighted magnetic resonance imaging scans in a total of 2095 patients with schizophrenia and 2861 healthy controls. Measures of grey matter volume and white matter microstructural integrity were extracted from the scans using harmonised protocols. For each measure, normative modelling was used to calculate how much patients deviated (in *z*-scores) from healthy controls at the individual level. Ordinal regression models were used to estimate the associations of these deviations with concurrent aggressive behaviour (as odds ratios [ORs] with 99% confidence intervals [CIs]). Mediation analyses were performed for positive symptoms (i.e., delusions, hallucinations and disorganised thinking), impulse control and illness insight. Aggression and potential mediators were assessed with the Positive and Negative Syndrome Scale, Scale for the Assessment of Positive Symptoms or Brief Psychiatric Rating Scale.

**Results:** Aggressive behaviour was significantly associated with reductions in total cortical volume (OR [99% CI] = 0.88 [0.78, 0.98], *p* = .003) and global white matter integrity (OR [99% CI] = 0.72 [0.59, 0.88], *p* = 3.50 × 10^−5^) and additional reductions in dorsolateral prefrontal cortex volume (OR [99% CI] = 0.85 [0.74, 0.97], *p* =.002), inferior parietal lobule volume (OR [99% CI] = 0.76 [0.66, 0.87], *p* = 2.20 × 10^−7^) and internal capsule integrity (OR [99% CI] = 0.76 [0.63, 0.92], *p* = 2.90 × 10^−4^). Except for inferior parietal lobule volume, these associations were largely mediated by increased severity of positive symptoms and reduced impulse control.

**Conclusions:** This study provides evidence that the co-occurrence of positive symptoms, poor impulse control and aggressive behaviour in schizophrenia has a neurobiological basis, which may inform the development of therapeutic interventions.

## 1. Introduction

Schizophrenia is associated with an increased risk of aggressive behaviour^1,2^. In a meta-analysis, the odds of engaging in violence, the most severe form of aggression^3^, were estimated to be approximately five times higher in individuals with schizophrenia than in the general population^1^. Important risk factors include substance misuse, poor impulse control, lack of illness insight and treatment nonadherence^4^. During a psychotic episode, aggressive behaviour may arise from positive symptoms – delusions, hallucinations or disorganised thinking^5^. However, the neurobiological correlates of aggression in schizophrenia remain poorly understood.

Brain regions typically implicated in aggressive behaviour are the orbitofrontal cortex, dorsolateral prefrontal cortex (DLPFC), anterior cingulate cortex, posterior cingulate cortex, insula, hippocampus, amygdala and striatum^6–8^. Recent systematic reviews of neuroimaging studies have pointed towards the precuneus and inferior parietal lobule (IPL) as being potentially important^9,10^. It has been hypothesised that reductions in the grey matter (GM) volume of these regions, or the microstructural integrity of the white matter (WM) tracts connected to them, lead to aggression via disturbances in emotion recognition and regulation, reward and avoidance learning and decision making^7^.

Schizophrenia is characterised by widespread reductions in GM volume and WM microstructural integrity^11^. The former are most pronounced in the frontal and temporal lobes, hippocampus, amygdala, nucleus accumbens and thalamus^12–14^ and the latter in interhemispheric, corticothalamic and frontotemporal pathways^15–17^. Furthermore, both are associated with increased symptom severity, cognitive deficits, poor illness insight and impaired social functioning^11,18^. The superior temporal gyrus, in particular, has been linked to positive symptoms^19^. Based on this, it may be hypothesised that illness-related reductions in GM volume and WM microstructural integrity underlie aggressive behaviour in schizophrenia.

To our knowledge, only one study has directly tested this hypothesis. This study, partly based on the same data as the current study, found that thinning of the anterior cingulate cortex, insula and inferior and middle temporal gyri in individuals with schizophrenia (*n* = 901) relative to healthy controls (*n* = 952) was associated with impulsive aggression across 10 sites worldwide^20^. However, limitations were the use of group-level differences between cases and controls as the exposure, the combination of impulsivity and aggression in the outcome measure and that subcortical regions and WM tracts were not investigated. A few studies have provided indirect tests by comparing aggressive and nonaggressive patients with healthy controls or by conducting analyses in patients only, with inconsistent results. Moreover, these studies have almost exclusively investigated GM and were limited by group-level analyses, small and selective samples of inpatients and potentially long time lags between brain scans and aggressive behaviour^21^.

To address the limitations of previous studies, we have investigated the associations of individual-level deviations from normative values of GM volume and WM microstructural integrity with concurrent aggressive behaviour in a large sample of individuals with schizophrenia from various care settings around the world.

## 2. Methods

### 2.1. Study samples

Data came from the Schizophrenia Working Group of the Enhancing Neuroimaging Genetics through Meta-Analysis (ENIGMA) consortium^22^. Twenty member sites contributed data to the current study. These sites were located in 13 different countries, with their samples totalling 2095 individuals diagnosed with schizophrenia (‘patients’) and 2861 unaffected general population controls (‘healthy controls’). Diagnoses were made in accordance with Diagnostic and Statistical Manual of Mental Disorders (DSM)-III^23^, DSM-IV^24^, DSM-5^25^ or the International Classification of Diseases (ICD)-10^26^. All participants gave written informed consent and the inclusion of each sample was approved by local ethics committees.

### 2.2. Image acquisition and processing

All 20 sites acquired high-resolution (voxel size of ≤1 mm^3^) T1-weighted magnetic resonance imaging scans and processed these using FreeSurfer^27^. Based on the Desikan-Killiany atlas^28^, structural measures were extracted for 34 cortical regions (cortical thickness [CT] and surface area [SA]) and 7 subcortical regions (volume) in each hemisphere. Nine sites also acquired diffusion-weighted images. From these images, values of fractional anisotropy (FA) and mean diffusivity (MD) were extracted for 25 WM tracts in each hemisphere. This was done using tract-based spatial statistics^29^ in conjunction with the John Hopkins University atlas^30^, implemented in FMRIB Software Library^31^. Additionally, sites extracted global (whole-brain) measures of CT, SA, FA and MD. The ENIGMA protocols (https://enigma.ini.usc.edu/protocols/), harmonised across sites, were followed for image processing and quality control. Information about the scanners, acquisition parameters and software versions used at the various sites has been published elsewhere^13,14,17^.

### 2.3. Variables

#### 2.3.1. Brain structure

We included all available global imaging-derived phenotypes (IPDs)^32^. In addition, we created a measure of total GM volume. This was calculated as the sum of total cortical volume (TCV) plus the combined volume of all subcortical regions. Cortical volumes were obtained by multiplying average CT by SA. A composite measure of WM microstructural integrity was created by averaging the patients’ deviation scores (see section 2.4.1) for FA and MD. Since greater integrity is assumed to be reflected by higher values of FA and lower values of MD^33^, we reversed (multiplied by -1) the deviation scores for the latter in the calculation of these averages.

Based on theory and previous studies, we included eight cortical (orbitofrontal cortex, DLPFC, anterior cingulate cortex, posterior cingulate cortex, insula, superior temporal gyrus, precuneus and IPL) and four subcortical (hippocampus, amygdala, striatum and thalamus) regions. The striatum, which FreeSurfer does not parcellate as a whole, was defined to encompass the caudate nucleus, putamen and nucleus accumbens^34^. For WM microstructure, we chose an exploratory approach because (i) research on the putative associations between WM abnormalities and aggression is lacking^9^ and (ii) nearly all major WM tracts in the cerebrum are connected to cortical or subcortical regions implicated in aggression^35^. We averaged values of FA or MD across segments of WM tracts where these had been extracted separately (e.g., the cingular and hippocampal parts of the cingulum). This yielded 14 WM tracts: corpus callosum, cingulum, fronto-occipital fasciculus, superior longitudinal fasciculus, sagittal stratum, uncinate fasciculus, corona radiata, internal capsule (IC), external capsule, corticospinal tract, posterior thalamic radiations, fornix and stria terminalis. With these, we included most of the major commissural, association and projection tracts in the cerebrum^35^.

#### 2.3.2. Aggressive behaviour

Depending on the site, aggressive behaviour was measured with item P7 (‘hostility’) of the Positive and Negative Syndrome Scale (PANSS)^36^, item B3 (‘aggressive and agitated behavior’) of the Scale for the Assessment of Positive Symptoms (SAPS)^37^ or item 10 (‘hostility’) of the Brief Psychiatric Rating Scale (BPRS)^38^ (Table S1). The PANSS was administered at most sites (*k* = 16), followed by the SAPS (*k* = 3) and BPRS (*k* = 1). Since items of the PANSS and BPRS are scored on a 7-point scale and those of the SAPS on a 6-point scale, we harmonised scores by collapsing the highest two option scores on the PANSS and BPRS. Given that scores of 7 (‘extreme’) on the PANSS (*n* = 2) and BPRS (*n* = 1) were rare, we did not think this approach detracted from the accuracy of our findings.

#### 2.3.3. Potential mediators and confounders

We included delusions, hallucinations, disorganised thinking, poor impulse control and lack of illness insight as potential mediators^39^. Table S1 shows which items of the PANSS, SAPS and BPRS were used for each. Scores were harmonised as described previously. Besides sex, age and global IDPs, we included three potential confounders: current dosage of antipsychotic medication (in chlorpromazine equivalents, calculated using the method by Woods^40^)^41^, duration of illness (DOI, defined as the number of years between the first psychotic episode and the scan date)^42^ and lifetime substance use disorder (SUD, based on DSM or ICD criteria)^39^.

### 2.4. Statistical analyses

#### 2.4.1. Normative modelling

For every IDP, we derived a normative model in the healthy controls using warped Bayesian linear regression with site, sex, age and – for local IDPs only – a global IDP as predictors. The predictors were selected for statistical (site) or theoretical (sex, age and global IDPs) reasons^43^. The global IDPs that served as predictors can be found in Table S2. The normative models were then run in the patients to calculate how much they deviated (in *z*-scores) from healthy controls on an individual level. Normative modelling was performed with the Predictive Clinical Neuroscience toolkit v0.25^44^.

#### 2.4.2. Primary analyses

We used one-sample *t*-tests to determine whether individuals with schizophrenia deviated – at the group level – from normative values (*z* = 0) of GM volume and WM microstructural integrity. Ordinal regression models were used to estimate the associations between such deviations and aggressive behaviour. Analyses were conducted for each IDP separately. We used cortical GM volume and the composite measure of WM microstructural integrity for the exposure variables. This was done to reduce the number of tests and facilitate comparison with previous studies^21^. Composite measures also capture the multidimensionality of brain morphology and may therefore be more sensitive to abnormalities therein than any of their components alone^45,46^. For reasons of statistical power, we chose TCV over total GM volume (missing for 458 more participants) as a variable of interest. We would expect negligible differences in point estimates based on these two measures, as TCV made up 90% of total GM volume on average. Values for local IDPs were averaged across hemispheres. To control for multiple testing, we set the testwise level of statistical significance at 1% instead of the more conventional 5%. The one-sample *t*-tests were run in SPSS v28.0^47^ and the ordinal regression models in Stata v17.0^48^.

#### 2.4.3. Mediation analyses

We performed a full mediation analysis (estimating paths Mx ⟹ Y, Xm ⟹ Y and X ⟹ M ⟹ Y) for an IDP if it was significantly (at the 1% level) associated with both a potential mediator and aggressive behaviour^49^. For the mediation analyses, we used generalised structural equation modelling in Stata v17.0^48^.

#### 2.4.4. Sensitivity analyses

We conducted three sets of sensitivity analyses. First, we examined the components of cortical GM volume (CT and SA) and WM microstructural integrity (FA and MD) individually. Second, we adjusted the analyses – one at a time – for current antipsychotic dosage, DOI and lifetime SUD. For this set of sensitivity analyses, the one-sample *t*-tests were replaced by ordinary least squares regression models with a forced intercept of 0 and a constant (xi = 1) as an independent variable (its coefficient being equal to the adjusted mean of deviation scores). Finally, we restricted the outcome to PANSS item P7 (thus leaving out SAPS item B3 and BPRS item 10).

## 3 Results

### 3.1. Sample characteristics

Tables 1 and 2 present the characteristics of participants by site and imaging modality. Overall, the mean age of patients was 35 years (SD = 12) and that of healthy controls was also 35 years (SD = 13). The proportion of men was higher in patients (*n* = 1391, 66%) than in healthy controls (*n* = 1404, 49%). Almost all patients were taking antipsychotic medication (*n* = 1675, 91%), with a median dosage equivalent to 300 mg of chlorpromazine (interquartile range [IQR] = 406). The median DOI was 7 years (IQR = 16). Of the patients for whom information about lifetime SUD was available (*n* = 778, 37%), almost half (*n* = 378, 42%) had such a diagnosis. Approximately one in seven patients displayed ‘mild’ or more severe aggression (*n* = 293, 15%) (Tables S3 and S4).

**Table 1.** Descriptive characteristics of participants who underwent T_1_-weighted magnetic resonance imaging.

| Site | SZ |  |  |  |  |  |  | HC |  |  |
| --- | --- | --- | --- | --- | --- | --- | --- | --- | --- | --- |
|  | *n* | Male, *n* (%) | Age, M (SD) | DOI, Mdn (IQR) | AP, *n* (%) | CPZ, Mdn (IQR) | SUD, *n* (%) | *n* | Male, *n* (%) | Age, M (SD) |
| COBRE | 67 | 55 (82) | 37 (14) | 11 (17) | 65 (98) | 300 (456) | - | 70 | 50 (71) | 36 (12) |
| ESO | 40 | 20 (50) | 29 (7) | - | 36 (100) | 332 (220) | - | 40 | 20 (50) | 29 (6) |
| FIDMAG | 124 | 95 (77) | 38 (11) | 13 (18) | 116 (99) | 450 (412) | - | 123 | 54 (44) | 38 (10) |
| FOR2107 | 124 | 63 (51) | 38 (12) | 15 (15) | 107 (86) | 300 (428) | 10 (8) | 944 | 344 (36) | 34 (13) |
| GAP | 53 | 10 (19) | 27 (6) | - | - | - | - | 94 | 57 (61) | 26 (7) |
| IGP | 68 | 40 (59) | 42 (11) | 19 (15) | 58 (87) | 200 (400) | - | 70 | 38 (54) | 36 (11) |
| IMH | 168 | 113 (67) | 34 (9) | 5 (8) | 168 (100) | 150 (150) | - | 110 | 66 (60) | 33 (10) |
| JBNU | 372 | 185 (50) | 38 (14) | 8 (16) | 285 (78) | 250 (507) | - | 358 | 173 (48) | 41 (14) |
| MCIC | 107 | 81 (76) | 34 (11) | 7 (18) | 97 (95) | 400 (600) | 67 (63) | 92 | 63 (68) | 33 (12) |
| Montreal | 87 | 87 (100) | 36 (10) | 13 (13) | 55 (100) | 589 (500) | 47 (54) | 40 | 40 (100) | 30 (7) |
| MPRC | 104 | 79 (76) | 34 (12) | - | - | - | - | 175 | 76 (43) | 37 (14) |
| NU | 105 | 72 (69) | 34 (13) | 5 (19) | 100 (95) | - | 63 (68) | 90 | 50 (56) | 32 (14) |
| Olin | 45 | 34 (76) | 29 (10) | 5 (6) | 38 (88) | 507 (439) | 16 (36) | 54 | 20 (37) | 35 (12) |
| RomeSL | 164 | 110 (67) | 39 (11) | 13 (17) | 153 (93) | 233 (267) | 33 (20) | 116 | 73 (63) | 38 (11) |
| RSCZ | 46 | 46 (100) | 22 (3) | 1 (1) | 38 (100) | - | - | 52 | 52 (100) | 22 (3) |
| SNUH | 39 | 18 (46) | 23 (6) | 5 (8) | 30 (77) | 142 (267) | - | 40 | 20 (50) | 23 (4) |
| UCISZ | 27 | 22 (81) | 43 (11) | 17 (16) | - | - | - | 30 | 23 (77) | 41 (12) |
| UMCU | 167 | 134 (80) | 27 (6) | 3 (5) | 149 (94) | 340 (375) | 78 (65) | 138 | 67 (49) | 27 (9) |
| UNINA | 45 | 31 (69) | 38 (10) | - | 45 (100) | 400 (393) | 10 (22) | 54 | 29 (54) | 43 (15) |
| Zürich | 60 | 45 (75) | 31 (8) | 7 (11) | 57 (95) | 333 (450) | - | 28 | 18 (64) | 33 (9) |
| *All* | *2012* | *1340 (67)* | *35 (12)* | *8 (16)* | *1597 (91)* | *320 (400)* | *324 (42)* | *2718* | *1333 (49)* | *35 (13)* |

SZ, individuals with schizophrenia; HC; healthy controls; M, mean; SD, standard deviation; DOI, duration of illness (in years); Mdn, median; IQR, interquartile range; AP, current antipsychotic use; CPZ, chlorpromazine equivalents (in mg); SUD, lifetime substance use disorder.

**Table 2.** Descriptive characteristics of participants who underwent diffusion-weighted magnetic resonance imaging.

| Site | SZ |  |  |  |  |  |  | HC |  |  |
| --- | --- | --- | --- | --- | --- | --- | --- | --- | --- | --- |
|  | *n* | Male, *n* (%) | Age, M (SD) | DOI, Mdn (IQR) | AP, *n* (%) | CPZ, Mdn (IQR) | SUD, *n* (%) | *n* | Male, *n* (%) | Age, M (SD) |
| COBRE | 55 | 43 (78) | 38 (14) | 12 (18) | 53 (98) | 300 (489) | - | 63 | 44 (70) | 35 (12) |
| ESO | 99 | 62 (63) | 30 (8) | - | 94 (96) | 332 (171) | - | 69 | 35 (51) | 27 (6) |
| FOR2107 | 104 | 53 (51) | 38 (11) | 15 (12) | 90 (87) | 300 (443) | 8 (8) | 856 | 300 (35) | 34 (13) |
| IMH | 170 | 115 (68) | 34 (9) | 5 (8) | 170 (100) | 150 (150) | - | 110 | 66 (60) | 33 (10) |
| RomeSL | 72 | 46 (64) | 40 (12) | 11 (18) | 67 (93) | 200 (300) | 13 (18) | 144 | 75 (52) | 48 (16) |
| RSCZ | 26 | 26 (100) | 22 (3) | 1 (1) | 24 (100) | - | - | 38 | 38 (100) | 22 (3) |
| SNUH | 39 | 18 (46) | 23 (6) | 5 (8) | 30 (77) | 142 (267) | - | 40 | 20 (50) | 23 (4) |
| UMCU | 124 | 103 (83) | 26 (6) | 3 (5) | 113 (94) | 383 (332) | 61 (69) | 87 | 42 (48) | 27 (8) |
| UNINA | 45 | 31 (69) | 38 (10) | - | 45 (100) | 400 (393) | 10 (22) | 54 | 29 (54) | 43 (15) |
| All | 734 | 497 (68) | 33 (11) | 6 (11) | 686 (94) | 252 (335) | 92 (30) | 1461 | 649 (44) | 34 (14) |

SZ, individuals with schizophrenia; HC; healthy controls; M, mean; SD, standard deviation; DOI, duration of illness (in years); Mdn, median; IQR, interquartile range; AP, current antipsychotic use; CPZ, chlorpromazine equivalents (in mg); SUD, lifetime substance use disorder.

### 3.2. Primary analyses

Reductions in TCV (odds ratio [OR] [99% CI] = 0.88 [0.78, 0.98], *p* = .003) (Table 3) and global WM microstructural integrity (OR [99% CI] = 0.72 [0.59, 0.88], *p* = 3.50 × 10^−5^) (Table 4) were significantly associated with aggressive behaviour. Associations between further region-specific reductions and aggression were found for DLPFC volume (OR [99% CI] = 0.85 [0.74, 0.97], *p* =.002), IPL volume (OR [99% CI] = 0.76 [0.66, 0.87], *p* = 2.20 × 10^−7^) and IC integrity (OR [99% CI] = 0.76 [0.63, 0.92], *p* = 2.90 × 10^−4^). Apart from IC integrity (*d* [99% CI] = 0.33 [0.22, 0.44], *p* = 1.98 × 10^−15^), all of these IDPs were reduced in patients compared with healthy controls (TCV: *d* [99% CI] = -0.46 [-0.52, -0.40], *p* = 3.59 × 10^−85^; global WM integrity: *d* [99% CI] = -0.28 [-0.39, -0.17], *p* = 1.03 × 10^−11^; DLPFC volume: *d* [99% CI] = -0.07 [-0.12, -0.01], *p* = .003; IPL volume: *d* [99% CI] = - 0.05 [-0.11, 0.01], *p* = .025). Relative sparing of GM volume or WM integrity was not associated with aggression.

**Table 3.** Deviations from normative values of grey matter volume (A) and their associations with aggressive behaviour (B) in individuals with schizophrenia (*N* = 2009).

| IDP | A |  |  | B |  |  |
| --- | --- | --- | --- | --- | --- | --- |
| | $d$ (99% CI) | SE | $p$ | OR (99% CI) | SE | $p$ |
| TCV | -0.46 (-0.52, -0.40) | 0.023 | $3.59 \times 10^{-85}$ | 0.88 (0.78, 0.98) | 0.039 | .003 |
| OFC | 0.06 (0.00, 0.11) | 0.022 | .014 | 1.08 (0.94, 1.23) | 0.054 | .151 |
| DLPFC | -0.07 (-0.12, -0.01) | 0.022 | .003 | 0.85 (0.74, 0.97) | 0.045 | .002 |
| ACC | -0.03 (-0.09, 0.02) | 0.022 | .142 | 0.99 (0.87, 1.13) | 0.051 | .835 |
| PCC | 0.08 (0.02, 0.13) | 0.022 | $7.95 \times 10^{-4}$ | 0.91 (0.80, 1.03) | 0.044 | .056 |
| INS | 0.08 (0.02, 0.14) | 0.022 | $3.50 \times 10^{-4}$ | 1.02 (0.89, 1.16) | 0.052 | .770 |
| STG | -0.09 (-0.15, -0.03) | 0.023 | $1.23 \times 10^{-4}$ | 1.11 (0.97, 1.26) | 0.056 | .044 |
| PREC | 0.12 (0.06, 0.17) | 0.022 | $2.43 \times 10^{-7}$ | 0.93 (0.82, 1.07) | 0.049 | .187 |
| IPL | -0.05 (-0.11, 0.01) | 0.023 | .025 | 0.76 (0.66, 0.87) | 0.040 | $2.20 \times 10^{-7}$ |
| HIPPO | -0.17 (-0.23, -0.11) | 0.023 | $7.94 \times 10^{-14}$ | 1.01 (0.89, 1.15) | 0.050 | .829 |
| AMYG | 0.06 (0.00, 0.12) | 0.023 | .007 | 1.02 (0.89, 1.16) | 0.052 | .754 |
| STRI | 0.37 (0.31, 0.43) | 0.024 | $6.19 \times 10^{-55}$ | 1.04 (0.93, 1.17) | 0.045 | .329 |
| THAL | -0.15 (-0.21, -0.09) | 0.024 | $3.82 \times 10^{-10}$ | 1.06 (0.93, 1.21) | 0.055 | .226 |

IDP, imaging-derived phenotype; CI, confidence interval; SE, standard error; OR, odds ratio; TCV, total cortical volume; OFC, orbitofrontal cortex; DLPFC, dorsolateral prefrontal cortex; ACC, anterior cingulate cortex; PCC, posterior cingulate cortex; INS, insula; STG, superior temporal gyrus; PREC, precuneus; IPL, inferior parietal lobule; HIPPO, hippocampus; AMYG, amygdala; STRI, striatum; THAL, thalamus.

**Table 4.** Deviations from normative values of white matter microstructural integrity (A) and their associations with aggressive behaviour (B) in individuals with schizophrenia (*N* = 608).

| IDP | A |  |  | B |  |  |
| --- | --- | --- | --- | --- | --- | --- |
| | $d$ (99% CI) | SE | $p$ | OR (99% CI) | SE | $p$ |
| GWM | -0.28 (-0.39, -0.17) | 0.041 | $1.03 \times 10^{-11}$ | 0.72 (0.59, 0.88) | 0.057 | $3.50 \times 10^{-5}$ |
| CC | -0.17 (-0.27, -0.06) | 0.041 | $4.60 \times 10^{-5}$ | 0.83 (0.66, 1.04) | 0.074 | .032 |
| CG | 0.33 (0.21, 0.46) | 0.049 | $1.13 \times 10^{-11}$ | 1.12 (0.87, 1.45) | 0.113 | .248 |
| FOF | 0.14 (0.03, 0.24) | 0.041 | $8.71 \times 10^{-4}$ | 0.98 (0.74, 1.29) | 0.106 | .825 |
| SLF | 0.11 (0.01, 0.22) | 0.041 | .005 | 1.14 (0.90, 1.44) | 0.105 | .154 |
| SS | 0.02 (-0.09, 0.12) | 0.041 | .651 | 1.10 (0.85, 1.42) | 0.111 | .352 |
| UNC | -0.07 (-0.18, 0.03) | 0.041 | .066 | 0.82 (0.63, 1.07) | 0.084 | .053 |
| CR | 0.02 (-0.08, 0.13) | 0.041 | .551 | 1.29 (0.99, 1.68) | 0.133 | .014 |
| IC | 0.33 (0.22, 0.44) | 0.042 | $1.98 \times 10^{-15}$ | 0.76 (0.63, 0.92) | 0.057 | $2.90 \times 10^{-4}$ |
| EC | 0.11 (0.01, 0.22) | 0.041 | .005 | 1.10 (0.85, 1.44) | 0.113 | .343 |
| CST | 0.11 (0.01, 0.22) | 0.041 | .007 | 1.09 (0.85, 1.40) | 0.106 | .376 |
| PTR | -0.05 (-0.15, 0.06) | 0.041 | .235 | 0.94 (0.71, 1.23) | 0.100 | .542 |
| FX | -0.23 (-0.34, -0.13) | 0.042 | $2.52 \times 10^{-8}$ | 1.09 (0.88, 1.35) | 0.090 | .279 |
| FXST | -0.05 (-0.15, 0.06) | 0.041 | .228 | 1.03 (0.80, 1.32) | 0.101 | .785 |

IDP, imaging-derived phenotype; CI, confidence interval; SE, standard error; OR, odds ratio; GWM, global white matter; CC, corpus callosum; CG, cingulum; FOF, fronto-occipital fasciculus; SLF, superior longitudinal fasciculus; SS, sagittal stratum; UNC, uncinate fasciculus; CR, corona radiata; IC, internal capsule; EC, external capsule; CST, corticospinal tract; PTR, posterior thalamic radiations; FX, fornix; FXST, fornix stria terminalis.

### 3.3. Mediation analyses

All potential mediators were significantly associated with at least one IDP and aggressive behaviour (Table 5). Delusions partly mediated the associations of TCV (OR [99% CI] = 0.94 [0.89, 0.98], *p* = 3.00 × 10^−4^), global WM integrity (OR [99% CI] = 0.83 [0.74, 0.94], *p* = 1.10 × 10^−4^) and DLPFC volume (OR [99% CI] = 0.93 [0.88, 0.98], *p* = 3.20 × 10^−4^) with aggressive behaviour, hallucinations those of TCV (OR [99% CI] = 0.96 [0.93, 0.99], *p* = 3.60 × 10^−4^) and global WM integrity (OR [99% CI] = 0.91 [0.84, 0.99], *p* = .003), disorganised thinking those of TCV (OR [99% CI] = 0.94 [0.90, 0.98], *p* = 3.70 × 10^−4^), global WM integrity (OR [99% CI] = 0.77 [0.66, 0.89], *p* = 5.50 × 10^−6^), DLPFC volume (OR [99% CI] = 0.92 [0.87, 0.97], *p* = 1.40 × 10^−5^) and IC integrity (OR [99% CI] = 0.81 [0.71, 0.94], *p* = 1.80 × 10^−4^) and poor impulse control those of TCV (OR [99% CI] = 0.86 [0.74, 0.99], *p* = .006), global WM integrity (OR [99% CI] = 0.70 [0.55, 0.88], *p* = 6.10 × 10^−5^), DLPFC volume (OR [99% CI] = 0.84 [0.72, 0.99], *p* = .006) and IC integrity (OR [99% CI] = 0.72 [0.58, 0.90], *p* = 1.60 × 10^−4^). None of the potential mediators was associated with IPL volume, so we did not perform mediation analyses for this IDP.

**Table 5.** Estimated paths between deviations in grey matter volume or white matter microstructural integrity (X), potential mediators (M) and aggressive behaviour (Y) in individuals with schizophrenia (*N* = 2095).

| IDP | $X \Rightarrow M$ | | | $M_x \Rightarrow Y$ | | | $X_m \Rightarrow Y$ | | | $X \Rightarrow M \Rightarrow Y$ | | |
| --- | --- | --- | --- | --- | --- | --- | --- | --- | --- | --- | --- | --- |
| | OR (99% CI) | SE | $p$ | OR (99% CI) | SE | $p$ | OR (99% CI) | SE | $p$ | OR (99% CI) | SE | $p$ |
| Delusions |  |  |  |  |  |  |  |  |  |  |  |  |
| TCV | 0.87 (0.79, 0.96) | 0.033 | $1.70 \times 10^{-4}$ | 1.63 (1.48, 1.80) | 0.064 | $3.90 \times 10^{-39}$ | 0.90 (0.79, 1.02) | 0.041 | .026 | 0.94 (0.89, 0.98) | 0.017 | $3.00 \times 10^{-4}$ |
| GWM | 0.76 (0.65, 0.90) | 0.045 | $3.00 \times 10^{-5}$ | 1.96 (1.66, 2.32) | 0.118 | $8.90 \times 10^{-25}$ | 0.81 (0.65, 1.01) | 0.061 | .014 | 0.83 (0.74, 0.94) | 0.037 | $1.10 \times 10^{-4}$ |
| DLPFC | 0.86 (0.77, 0.95) | 0.033 | $1.80 \times 10^{-4}$ | 1.64 (1.48, 1.80) | 0.059 | $2.10 \times 10^{-38}$ | 0.90 (0.78, 1.04) | 0.047 | .066 | 0.93 (0.88, 0.98) | 0.019 | $3.20 \times 10^{-4}$ |
| Hallucinations |  |  |  |  |  |  |  |  |  |  |  |  |
| TCV | 0.87 (0.79, 0.95) | 0.030 | $9.80 \times 10^{-5}$ | 1.33 (1.22, 1.44) | 0.041 | $8.30 \times 10^{-19}$ | 0.90 (0.80, 1.01) | 0.039 | .021 | 0.96 (0.93, 0.99) | 0.011 | $3.60 \times 10^{-4}$ |
| GWM | 0.80 (0.67, 0.95) | 0.049 | $8.60 \times 10^{-4}$ | 1.51 (1.29, 1.77) | 0.086 | $2.10 \times 10^{-11}$ | 0.76 (0.62, 0.94) | 0.056 | $8.10 \times 10^{-4}$ | 0.91 (0.84, 0.99) | 0.027 | .003 |
| Disorganised thinking |  |  |  |  |  |  |  |  |  |  |  |  |
| TCV | 0.87 (0.79, 0.96) | 0.031 | $1.70 \times 10^{-4}$ | 1.51 (1.37, 1.66) | 0.054 | $9.30 \times 10^{-28}$ | 0.90 (0.80, 1.02) | 0.039 | .028 | 0.94 (0.90, 0.98) | 0.015 | $3.70 \times 10^{-4}$ |
| GWM | 0.70 (0.58, 0.84) | 0.045 | $3.00 \times 10^{-7}$ | 2.08 (1.71, 2.52) | 0.141 | $1.00 \times 10^{-22}$ | 0.82 (0.66, 1.02) | 0.062 | .020 | 0.77 (0.66, 0.89) | 0.041 | $5.50 \times 10^{-6}$ |
| DLPFC | 0.81 (0.73, 0.91) | 0.034 | $1.90 \times 10^{-6}$ | 1.50 (1.36, 1.65) | 0.054 | $1.40 \times 10^{-26}$ | 0.89 (0.78, 1.02) | 0.045 | .032 | 0.92 (0.87, 0.97) | 0.017 | $1.40 \times 10^{-5}$ |
| IC | 0.76 (0.64, 0.91) | 0.047 | $5.70 \times 10^{-5}$ | 2.15 (1.77, 2.60) | 0.146 | $1.50 \times 10^{-24}$ | 0.79 (0.64, 0.97) | 0.057 | .003 | 0.81 (0.71, 0.94) | 0.042 | $1.80 \times 10^{-4}$ |
| Poor impulse control |  |  |  |  |  |  |  |  |  |  |  |  |
| TCV | 0.87 (0.76, 0.99) | 0.041 | .005 | 2.98 (2.56, 3.47) | 0.163 | $1.30 \times 10^{-76}$ | 0.87 (0.76, 1.01) | 0.045 | .013 | 0.86 (0.74, 0.99) | 0.045 | .006 |
| GWM | 0.70 (0.57, 0.86) | 0.052 | $1.30 \times 10^{-5}$ | 2.77 (2.14, 3.59) | 0.245 | $3.20 \times 10^{-24}$ | 0.91 (0.73, 1.13) | 0.069 | .270 | 0.70 (0.55, 0.88) | 0.056 | $6.10 \times 10^{-5}$ |
| DLPFC | 0.85 (0.73, 0.99) | 0.046 | $5.60 \times 10^{-3}$ | 2.96 (2.54, 3.45) | 0.163 | $2.00 \times 10^{-74}$ | 0.89 (0.76, 1.05) | 0.051 | .061 | 0.84 (0.72, 0.99) | 0.049 | .006 |
| IC | 0.72 (0.59, 0.89) | 0.052 | $4.60 \times 10^{-5}$ | 2.73 (2.11, 3.52) | 0.239 | $5.50 \times 10^{-24}$ | 0.81 (0.65, 1.00) | 0.060 | $8.40 \times 10^{-3}$ | 0.72 (0.58, 0.90) | 0.056 | $1.60 \times 10^{-4}$ |
| Lack of illness insight |  |  |  |  |  |  |  |  |  |  |  |  |
| DLPFC | 0.87 (0.77, 1.00) | 0.042 | $9.50 \times 10^{-3}$ | 1.61 (1.42, 1.83) | 0.074 | $3.00 \times 10^{-22}$ | 0.93 (0.77, 1.13) | 0.063 | .360 | 0.94 (0.88, 1.00) | 0.023 | .012 |

IDP, imaging-derived phenotype; OR, odds ratio; CI, confidence interval; SE, standard error; TCV, total cortical volume; GWM, global white matter; DLPFC, dorsolateral prefrontal cortex; IC, internal capsule.

### 3.4. Sensitivity analyses

The association of global WM microstructural integrity with aggressive behaviour was driven by both FA and MD, whereas those of TCV, DLPFC volume and IC integrity were mainly driven by either SA or FA (Tables S5-S8). Significant associations between IDPs and aggression remained so after adjustment for current antipsychotic dosage (Tables S9 and S10), DOI (Tables S11 and S12) or – with the exception of IC integrity – lifetime SUD (Tables S13 and S14). Similar results were obtained when using only PANSS item P7 as the outcome (Tables S15 and S16).

## 4 Discussion

The findings of this study are consistent with the hypothesis that illness-related changes in brain structure, consisting of widespread reductions in cortical GM volume and WM microstructural integrity, underlie aggressive behaviour in schizophrenia^39^. Furthermore, they suggest that these changes may lead to aggression via increased occurrence or severity of positive symptoms – delusions, hallucinations and disorganised thinking – and reduced impulse control. Delusions and hallucinations may provide motivation for aggression^50^, especially if they are accompanied by anger^51,52^ or poor illness insight^53^. Disorganised thinking mostly reflects impairments in executive functions^54^, which have been linked to aggressive behaviour in schizophrenia^55,56^. Depending on what executive function is impaired (in parentheses), aggression may result from a diminished ability to supress prepotent responses (response inhibition)^8^, change behaviour in the face of negative consequences (cognitive flexibility)^57^, anticipate the possible consequences thereof (planning)^58^ or find a more adaptive solution in provocative situations (reasoning and problem solving)^59^. Impulsivity is the nonreflective selection of a stimulus-evoked response or an immediately rewarding response^60^ and, as such, lowers the threshold for aggression.

The effects of brain structure on aggressive behaviour in schizophrenia appear to be global rather than regional. This corresponds to reports of global reductions in cortical GM volume^61,62^ and WM microstructural integrity^63^ in individuals with antisocial personality disorder (ASPD) or psychopathy, two psychiatric disorders characterised by lifelong antisocial – and often aggressive – behaviour^64^. However, we found additive effects for the DLPFC, IPL and IC. The DLPFC plays a central role in cognitive control, which is the ability to hold relevant information in working memory when guiding behaviour towards a goal^65^. Cognitive control subserves higher-order executive functions, namely planning, reasoning and problem solving, and impulse control^66^. In line with this, reduced volume or activation of the DLPFC has consistently been found to be associated with executive dysfunction^67,68^ and impulsivity^69^ in schizophrenia. Executive functions contribute to illness insight^70^. This may explain why poor illness insight and delusions, whose definition requires the former^71^, mediated the association between DLPFC volume and aggression. The IPL is centrally involved in theory of mind, or the ability to infer mental states in oneself and others^72^. Impaired theory of mind is a core feature of schizophrenia^73^ and increases the risk of aggressive behaviour^55,74,75^. It may do so through misinterpretation of social cues^76^, difficulties with identifying or understanding emotions^77^, blurring of self-other boundaries^76^ or a lack of empathy^78^. Since we did not test for theory of mind, this is an important avenue for future research. The IC, as the point of convergence for nerve tracts connecting the prefrontal cortex with subcortical regions, is part of structural brain networks that modulate a wide range of cognitive processes^79^. Accordingly, damage to the IC has long been hypothesised to be a cause of cognitive impairment in schizophrenia^80,81^. Cognitive processes that the IC facilitates and, when impaired, may promote aggressive behaviour are response inhibition, memory formation, emotion regulation and reward sensitivity^79^. As mentioned, response inhibition and reward sensitivity are determinants of impulsivity. Memory formation is necessary to learn from the consequences of one’s actions^82^. Emotional dysregulation means that one may act on or attempt to alleviate negative emotions, such as anger, fear and frustration, with aggression when this is not situationally appropriate^83^.

### 4.1. Strengths and limitations

To our knowledge, this is the largest study to date investigating the relationship between structural brain abnormalities and aggressive behaviour in schizophrenia. It is also the first in this field of research to measure such abnormalities on an individual level. Other strengths include the representativeness of the sample and the wide range of tests for robustness and underlying mechanisms. However, our study has a number of important limitations. First, its cross-sectional design precludes causal inference. However, scanning was done in close temporal proximity to the measurement of aggression and we adjusted for several potential confounders. Second, aggressive behaviour was only measured at one point in time. This may have attenuated effects, as brain abnormalities are likely more pronounced in patients with recurrent rather than incidental aggression^7^. In addition, acutely agitated patients are difficult to engage in research^84^ and scan successfully^85^. However, such patients would have been invited to participate once they were clinically stable. Third, aggressive behaviour was measured with individual items of instruments designed for measuring symptoms and clinical features of schizophrenia. Although these items are commonly used as outcome measures^4^, multi-item instruments purposely designed for measuring aggression will be more sensitive. Fourth, aggressive behaviour and potential mediators were measured with the same instruments. Inter-item correlations may therefore have contributed to the mediation effects, despite items measuring different constructs. Fifth, there was no information about comorbid ASPD or psychopathy. The structural brain correlates of aggression may differ between patients with and without such comorbidity. However, most (an estimated 75%) aggressive behaviour in schizophrenia begins at or after first-episode psychosis^86^ and is, for that reason, unlikely related to ASPD or psychopathy. Finally, we investigated the pairwise associations between brain regions and aggression. While many cortical and subcortical regions are functionally specialised, complex behaviours arise from their interaction in networks^87^. It is possible that these networks exert a stronger effect on aggression than any brain region on its own.

### 4.2. Clinical implications and recommendations for future research

The findings of this study may further the development of prevention strategies for aggressive behaviour in individuals with schizophrenia in that they provide: (i) candidate neuroimaging markers for predicting treatment response; (ii) potential targets for neuromodulation therapies, such as deep brain stimulation and transcranial magnetic stimulation; and (iii) support for the use of other nonpharmacological interventions, in particular cognitive behavioural therapy, cognitive remediation therapy and physical exercise, that preliminary evidence suggests have neuroprotective or neurogenic effects^88,89^. The findings also provide a rationale for investigating whether structural neuroimaging adds to the limited^90,91^ predictive performance of current risk assessment instruments. However, benefit-harm balance and cost-effectiveness would be important considerations, as the observed effects were modest and largely mediated by risk factors that are more amenable to change and less expensive to measure than brain structure. To clarify causal mechanisms, we recommend that future studies use longitudinal designs and test for specific cognitive functions as potential mediators. Network analysis^92^ may improve our understanding of the structural brain networks that underlie aggression in schizophrenia.

## Data Availability

Data for this study are not available, as the participants did not agree for these to be shared publicly.

## Financial support

The ENIGMA project is supported by the National Institute of Biomedical Imaging and Bioengineering of the National Institutes of Health (NIH) under grant number U54EB020403. Funding information for the participating sites can be found in the supplement. AR is supported by the Richard Perry University Professorship, and TN-J by the Andrew H. Woods Professorship. The funders had no role in study design, data collection or analysis, decision to publish or the preparation of the manuscript.

## Notes

### Competing Interest Statement

The authors have declared no competing interest.

### Author Declarations

This work was approved by ethics committees at each site.

