## Supplementary materials for "Structural brain abnormalities and aggressive behaviour in schizophrenia: Mega-analysis of data from 2095 patients and 2861 healthy controls via the ENIGMA consortium"

to:

### Funding information for participating ENIGMA sites

**COBRE**
The COBRE study was supported by the National Institutes of Health (NIH) (grants R01EB006841 and P20GM103472) and National Science Foundation (NSF) (grant 1539067). JAT and VDC are supported by NIH grants 5R01MH094524 and 5R01MH121246.

**ESO**
The ESO study was supported by the Ministry of Health of the Czech Republic (grants NU21-08-00432 and NU22-04-00143).

**FIDMAG**
The FIDMAG study was supported by a Miguel Servet Research Contract (MS14/00041), the Plan Nacional de I+D+i 2013–2016 (Research Project PI14/00292), a Juan de la Cierva-formación contract (FJCI-2015-25278), the Instituto de Salud Carlos III-Subdirección General de Evaluación y Fomento de la Investigación, the European Regional Development Fund and the Centro de Investigación Biomédica en Red de Salud Mental.

**FOR2107**
The FOR2107 study was supported by grants from the German Research Foundation to TK (KI 588/14-1 and KI 588/14-2), UD (DA 1151/5-1, DA 1151/5-2, DA1151/9-1, DA1151/10-1 and DA1151/11-1) and IN (NE2254/1-2, NE2254/2-1, NE2254/3-1 and NE2254/4-1). UD is also supported by the Interdisciplinary Centre for Clinical Research of the University of Münster (grant Dan3/022/22).

**GAP**
The GAP study was supported by the NIHR Biomedical Research Centre at South London and Maudsley NHS Foundation Trust and King’s College London.

**IGP**
The IGP study was supported by grants (APP630471 and APP1081603) and a Career Development Award (APP1061875) from the Australian National Health and Medical Research Council and a grant (CE110001021) from the Australian Research Council Centre of Excellence in Cognition and its Disorders to MJG.

**IMH**
This work was supported by grants from the National Healthcare Group (SIG/05004 and SIG/05028) and the Singapore Bioimaging Consortium (RP C009/2006) to KS.

**JBNU**
This work was supported by grants from the Korean Mental Health Technology R&D Project (HL19C0015) and the Korea Health Industry Development Institute (HR18C0016) to Y-CC.

**MCIC**
The MCIC study was supported by the NIH (grants P41RR14075 and R01EB005846 to VDC), Department of Energy (DE-FG02-99ER62764), Mind Research Network, Morphometry (1U24, RR021382A) and Function (U24RR021992-01, NIH.NCRR MO1 RR025758-01 and NIMH 1RC1MH089257 to VDC) testbeds of the Biomedical Informatics Research Network, German Research Foundation (research fellowship to SE), National Alliance for Research on Schizophrenia & Depression (Young Investigator Award to SE) and Else-Kröner Fresenius Foundation (grant 2019-A118 to DA).

**Montreal**
This work was supported through the Eli Lilly Chair of Schizophrenia at the University of Montreal, held by SP.

**MPRC**
The MPRC study was supported by NIH grants (U01MH108148, 2R01EB015611, R01MH112180, R01DA027680, R01MH085646, P50MH103222 and T32MH067533), a State of Maryland contract (M00B6400091) and an NSF grant (1620457).

**NU**
The NU study was supported by the NIH (grants P50 MH071616, R01 EB020062, R01 MH056584, R01 MH084803 and U01 MH097435 to LW and grant T32 NS047987 to DC) and NSF (grants 1636893 and NSF 1734853 to LW).

**Olin**
The Olin study was supported by NIH grant R01MH077945.

**RomeSL**
This work was supported by the Italian Ministry of Health (Ricerca Corrente 2023).

**RSCZ**
The RSCZ study was supported by the Russian Foundation for Basic Research (grant 20-013-00748).

**SNUH**
This work was supported by the National Research Foundation of Korea and Korea Brain Research Institute (grants 2019R1C1C1002457, 2020M3E5D9079910, RS-2023-00266120 and 21-BR-03-01).

**UCISZ**
The UCISZ study was supported by the National Institutes of Mental Health (grant R21MH097196 to TGMvE). Data were processed with the high-performance computing cluster at the University of California, Irvine, supported by Joseph Farran, Harry Mangalam, Adam Brenner, the National Center for Research Resources and National Center for Advancing Translational Sciences (grant UL1 TR000153).

**UMCU**
This work was supported by the Geestkracht programme of the Dutch Health Research Council (grant 10-000-1001), University Medical Center Utrecht and participating pharmaceutical companies (Lundbeck, AstraZeneca, Eli Lilly, Janssen Cilag) and mental healthcare institutions (Altrecht, GGZ Centraal and Delta).

**UNINA**
FI, AdB, MC and AB were supported by NextGenerationEU, the Italian Ministry of Education, University and Research, National Recovery and Resilience Plan and project MNESYS (PE0000006) – A Multiscale integrated approach to the study of the nervous system in health and disease (DN. 1553 11.10.2022).

**Zürich**
This work was supported by the Swiss National Science Foundation.

**Table S1.** Items used to measure aggressive behaviour and potential mediators.

| Variable | Item |
|---|---|
| Aggressive behaviour | PANSS item P7 'hostility' |
| | SAPS item B3 'aggressive and agitated behaviour' |
| | BPRS item 10 'hostility' |
| Delusions | PANSS item P1 'delusions' |
| | SAPS item D13 'global rating of severity of delusions' |
| Hallucinations | PANSS item P3 'hallucinatory behaviour' |
| | SAPS item H7 'global rating of severity of hallucinations' |
| | BPRS item 12 'hallucinatory behaviour' |
| Disorganised thinking | PANSS item P2 'conceptual disorganisation' |
| | SAPS item P9 'global rating of positive formal thought disorder' |
| | BPRS item 4 'conceptual disorganisation' |
| Poor impulse control | PANSS item P14 'poor impulse control' |
| Lack of illness insight | PANSS item G12 'lack of judgment and insight' |

PANSS, Positive and Negative Syndrome Scale; SAPS, Scale for the Assessment of Positive Symptoms; BPRS, Brief Psychiatric Rating Scale.

**Table S2.** Global imaging-derived phenotypes used in the normative models as predictors of local imaging-derived phenotypes.

| Class of local IDPs | Global IDP |
|---|---|
| GM volume | Total cortical volume (cortical regions) |
| | Total GM volume (subcortical regions) |
| Cortical thickness | Average CT across the entire cortex |
| Surface area | Total SA of the cortex |
| WM microstructural integrity | Average of deviation scores for global FA and global MD |
| Fractional anisotropy | Average FA across the entire WM skeleton |
| Mean diffusivity | Average MD across the entire WM skeleton |

IDP, imaging-derived phenotype; GM, grey matter; CT, cortical thickness; SA, surface area; WM, white matter; FA, fractional anisotropy; MD, mean diffusivity.

**Table S3.** Distribution of aggression scores in individuals with schizophrenia who underwent $T_1$-weighted magnetic resonance imaging.

| | Item | | | |
|---|---|---|---|---|
| **Score** | PANSS P7 (*n* = 1572) | SAPS B3 (*n* = 336) | BPRS 10 (*n* = 104) | Total (*N* = 2012) |
| 1 'none' | 1165 (74) | 280 (83) | 63 (61) | 1508 (75) |
| 2 'minimal' | 163 (10) | 25 (7) | 23 (22) | 211 (10) |
| 3 'mild' | 150 (10) | 22 (7) | 13 (13) | 185 (9) |
| 4 'moderate' | 65 (4) | 6 (2) | 2 (2) | 73 (4) |
| 5 'marked' | 22 (1) | 2 (<1) | 1 (1) | 25 (1) |
| 6 'severe' | 7 (<1) | 1 (<1) | 2 (2) | 10 (<1) |

Data are *n* (%). PANSS, Positive and Negative Syndrome Scale; SAPS, Scale for the Assessment of Positive Symptoms; BPRS, Brief Psychiatric Rating Scale.

**Table S4.** Distribution of aggression scores in individuals with schizophrenia who underwent diffusion-weighted magnetic resonance imaging.

| | Item | | |
|---|---|---|---|
| **Score** | PANSS P7 (*n* = 630) | SAPS B3 (*n* = 100) | Total (*N* = 734) |
| 1 'none' | 441 (70) | 98 (94) | 539 (73) |
| 2 'minimal' | 83 (13) | 3 (3) | 86 (12) |
| 3 'mild' | 56 (9) | 3 (3) | 59 (8) |
| 4 'moderate' | 31 (5) | - | 31 (4) |
| 5 'marked' | 14 (2) | - | 14 (2) |
| 6 'severe' | 5 (<1) | - | 5 (<1) |

Data are *n* (%). PANSS, Positive and Negative Syndrome Scale; SAPS, Scale for the Assessment of Positive Symptoms.

**Table S5.** Deviations from normative values of cortical thickness (A) and their associations with aggressive behaviour (B) in individuals with schizophrenia (*N* = 2012).

| | A | | | B | | |
|---|---|---|---|---|---|---|
| **IDP** | *d* (99% CI) | SE | *p* | OR (99% CI) | SE | *p* |
| GCT | -0.63 (-0.70, -0.56) | 0.024 | $4.24 \times 10^{-147}$ | 1.03 (0.91, 1.16) | 0.047 | .571 |
| OFC | 0.00 (-0.06, 0.06) | 0.022 | .905 | 1.22 (1.07, 1.39) | 0.061 | $1.00 \times 10^{-4}$ |
| DLPFC | -0.09 (-0.14, -0.03) | 0.022 | $1.37 \times 10^{-4}$ | 0.95 (0.82, 1.08) | 0.050 | .295 |
| ACC | 0.15 (0.10, 0.21) | 0.023 | $9.16 \times 10^{-12}$ | 1.13 (0.99, 1.28) | 0.057 | .016 |
| PCC | 0.05 (-0.01, 0.10) | 0.022 | .042 | 1.03 (0.90, 1.17) | 0.052 | .603 |
| INS | -0.05 (-0.11, 0.01) | 0.022 | .023 | 1.02 (0.90, 1.16) | 0.051 | .684 |
| STG | -0.09 (-0.15, -0.03) | 0.023 | $1.41 \times 10^{-4}$ | 1.07 (0.94, 1.21) | 0.052 | .183 |
| PREC | 0.14 (0.08, 0.19) | 0.022 | $1.04 \times 10^{-9}$ | 0.91 (0.80, 1.04) | 0.046 | .060 |
| IPL | 0.02 (-0.03, 0.08) | 0.022 | .303 | 0.85 (0.75, 0.97) | 0.042 | .001 |

IDP, imaging-derived phenotype; CI, confidence interval; SE, standard error; OR, odds ratio; GCT, global cortical thickness; OFC, orbitofrontal cortex; DLPFC, dorsolateral prefrontal cortex; ACC, anterior cingulate cortex; PCC, posterior cingulate cortex; INS, insula; STG, superior temporal gyrus; PREC, precuneus; IPL, inferior parietal lobule.

**Table S6.** Deviations from normative values of surface area (A) and their associations with aggressive behaviour (B) in individuals with schizophrenia (*N* = 2009).

| | A | | | B | | |
|---|---|---|---|---|---|---|
| **IDP** | *d* (99% CI) | SE | *p* | OR (99% CI) | SE | *p* |
| GSA | -0.21 (-0.26, 0.15) | 0.023 | $8.75 \times 10^{-20}$ | 0.85 (0.76, 0.95) | 0.039 | $3.20 \times 10^{-4}$ |
| OFC | 0.06 (0.01, 0.12) | 0.022 | .004 | 0.97 (0.86, 1.09) | 0.046 | .499 |
| DLPFC | -0.05 (-0.10, .01) | 0022 | .042 | 0.88 (0.77, 1.00) | 0.044 | .008 |
| ACC | -0.13 (-0.19, -0.08) | 0.023 | $2.45 \times 10^{-9}$ | 0.97 (0.85, 1.10) | 0.050 | .493 |
| PCC | -0.03 (-0.08, 0.03) | 0.022 | .240 | 0.94 (0.83, 1.06) | 0.046 | .198 |
| INS | 0.04 (-0.02, 0.09) | 0.022 | .108 | 1.01 (0.89, 1.16) | 0.052 | .795 |
| STG | 0.09 (0.03, 0.15) | 0.023 | $7.40 \times 10^{-5}$ | 1.17 (1.05, 1.30) | 0.050 | $2.30 \times 10^{-4}$ |
| PREC | 0.08 (0.03, 0.14) | 0.022 | $1.64 \times 10^{-4}$ | 0.97 (0.85, 1.10) | 0.050 | .510 |
| IPL | -0.04 (-0.10, 0.02) | 0.023 | .068 | 0.82 (0.72, 0.93) | 0.042 | $9.00 \times 10^{-5}$ |

IDP, imaging-derived phenotype; CI, confidence interval; SE, standard error; OR, odds ratio; GSA, global surface area; OFC, orbitofrontal cortex; DLPFC, dorsolateral prefrontal cortex; ACC, anterior cingulate cortex; PCC, posterior cingulate cortex; INS, insula; STG, superior temporal gyrus; PREC, precuneus; IPL, inferior parietal lobule.

**Table S7.** Deviations from normative values of fractional anisotropy (A) and their associations with aggressive behaviour (B) in individuals with schizophrenia (*N* = 733).

| | A | | | B | | |
|---|---|---|---|---|---|---|
| **IDP** | *d* (99% CI) | SE | *p* | OR (99% CI) | SE | *p* |
| GFA | -0.18 (-0.27, -0.08) | 0.037 | $2.00 \times 10^{-6}$ | 0.82 (0.69, 0.97) | 0.054 | .003 |
| CC | -0.17 (-0.27, -0.07) | 0.037 | $5.00 \times 10^{-6}$ | 0.88 (0.73, 1.06) | 0.065 | .083 |
| CG | 0.21 (0.10, 0.32) | 0.043 | $1.00 \times 10^{-6}$ | 0.97 (0.79, 1.19) | 0.079 | .691 |
| FOF | 0.02 (-0.08, 0.11) | 0.037 | .603 | 0.97 (0.79, 1.20) | 0.080 | .713 |
| SLF | 0.13 (0.03, 0.23) | 0.037 | $4.47 \times 10^{-4}$ | 1.12 (0.92, 1.35) | 0.083 | .145 |
| SS | -0.05 (-0.15, 0.04) | 0.037 | .152 | 0.90 (0.73, 1.10) | 0.070 | .171 |
| UNC | 0.05 (-0.04, 0.15) | 0.037 | .150 | 1.05 (0.84, 1.31) | 0.090 | .576 |
| CR | 0.07 (-0.03, 0.16) | 0.037 | .060 | 1.27 (1.06, 1.53) | 0.092 | .001 |
| IC | 0.36 (0.26, 0.46) | 0.038 | $1.86 \times 10^{-21}$ | 0.81 (0.70, 0.94) | 0.047 | $2.80 \times 10^{-4}$ |
| EC | 0.12 (0.02, 0.21) | 0.037 | .001 | 1.15 (0.94, 1.41) | 0.090 | .075 |
| CST | -0.03 (-0.12, 0.07) | 0.037 | .434 | 0.99 (0.82, 1.20) | 0.074 | .903 |
| PTR | -0.01 (-0.10, 0.09) | 0.037 | .828 | 1.01 (0.84, 1.23) | 0.076 | .845 |
| FX | -0.12 (-0.22, -0.02) | 0.037 | .001 | 1.01 (0.85, 1.19) | 0.065 | .896 |
| FXST | -0.05 (-0.15, 0.05) | 0.037 | .175 | 1.10 (0.90, 1.35) | 0.086 | .212 |

IDP, imaging-derived phenotype; CI, confidence interval; SE, standard error; OR, odds ratio; GFA, global fractional anisotropy; CC, corpus callosum; CG, cingulum; FOF, fronto-occipital fasciculus; SLF, superior longitudinal fasciculus; SS, sagittal stratum; UNC, uncinate fasciculus; CR, corona radiata; IC, internal capsule; EC, external capsule; CST, corticospinal tract; PTR, posterior thalamic radiations; FX, fornix; FXST, fornix stria terminalis.

**Table S8.** Deviations from normative values of mean diffusivity (A) and their associations with aggressive behaviour (B) in individuals with schizophrenia (*N* = 607).

| | A | | | B | | |
|---|---|---|---|---|---|---|
| **IDP** | *d* (99% CI) | SE | *p* | OR (99% CI) | SE | *p* |
| GMD | 0.27 (0.16, 0.37) | 0.041 | $1.04 \times 10^{-10}$ | 1.41 (1.18, 1.68) | 0.097 | $8.30 \times 10^{-7}$ |
| CC | 0.10 (0.00, 0.21) | 0.041 | .011 | 1.13 (0.94, 1.35) | 0.079 | .086 |
| CG | -0.15 (-0.26, -0.05) | 0.041 | $2.25 \times 10^{-4}$ | 0.78 (0.64, 0.93) | 0.056 | $3.80 \times 10^{-4}$ |
| FOF | -0.15 (-0.25, -0.04) | 0.041 | $3.40 \times 10^{-4}$ | 1.00 (0.81, 1.23) | 0.080 | .958 |
| SLF | -0.04 (-0.15, 0.06) | 0.041 | .294 | 0.94 (0.78, 1.13) | 0.068 | .408 |
| SS | -0.08 (-0.18, 0.03) | 0.041 | .061 | 0.82 (0.67, 1.00) | 0.063 | .010 |
| UNC | 0.12 (0.02, 0.23) | 0.041 | .002 | 1.23 (1.01, 1.49) | 0.093 | .007 |
| CR | 0.04 (-0.07, 0.14) | 0.041 | .381 | 0.95 (0.76, 1.18) | 0.081 | .560 |
| IC | -0.09 (-0.19, 0.02) | 0.041 | .029 | 1.10 (0.93, 1.31) | 0.072 | .133 |
| EC | -0.09 (-0.19, 0.02) | 0.041 | .034 | 1.02 (0.83, 1.25) | 0.081 | .841 |
| CST | -0.19 (-0.30, -0.09) | 0.041 | $2.00 \times 10^{-6}$ | 0.91 (0.76, 1.10) | 0.067 | .212 |
| PTR | 0.05 (-0.06, 0.15) | 0.041 | .246 | 1.04 (0.86, 1.27) | 0.079 | .582 |
| FX | 0.24 (0.13, 0.35) | 0.042 | $1.09 \times 10^{-8}$ | 0.94 (0.77, 1.13) | 0.070 | .378 |
| FXST | 0.01 (-0.09, 0.11) | 0.041 | .809 | 1.09 (0.92, 1.29) | 0.072 | .188 |

IDP, imaging-derived phenotype; CI, confidence interval; SE, standard error; OR, odds ratio; GMD, global mean diffusivity; CC, corpus callosum; CG, cingulum; FOF, fronto-occipital fasciculus; SLF, superior longitudinal fasciculus; SS, sagittal stratum; UNC, uncinate fasciculus; CR, corona radiata; IC, internal capsule; EC, external capsule; CST, corticospinal tract; PTR, posterior thalamic radiations; FX, fornix; FXST, fornix stria terminalis.

**Table S9.** Deviations from normative values of grey matter volume (A) and their associations with aggressive behaviour (B) in individuals with schizophrenia, adjusting for current dosage of antipsychotic medication ($N$ = 1499).

| | A | | | B | | |
|---|---|---|---|---|---|---|
| **IDP** | *d* (99% CI) | SE | *p* | OR (99% CI) | SE | *p* |
| TCV | -0.33 (-0.40, -0.26) | 0.026 | $1.67 \times 10^{-35}$ | 0.84 (0.73, 0.96) | 0.046 | .001 |
| OFC | 0.06 (-0.01, 0.12) | 0.026 | .029 | 1.09 (0.94, 1.28) | 0.066 | .136 |
| DLPFC | -0.03 (-0.09, 0.04) | 0.026 | .326 | 0.85 (0.73, 1.00) | 0.052 | .009 |
| ACC | -0.02 (-0.08, 0.05) | 0.026 | .519 | 0.98 (0.84, 1.14) | 0.058 | .715 |
| PCC | 0.06 (-0.01, 0.12) | 0.026 | .028 | 0.89 (0.77, 1.03) | 0.050 | .038 |
| INS | 0.07 (0.00, 0.14) | 0.026 | .008 | 1.06 (0.91, 1.24) | 0.063 | .325 |
| STG | -0.06 (-0.13, 0.01) | 0.027 | .033 | 1.18 (1.02, 1.38) | 0.069 | .004 |
| PREC | 0.12 (0.05, 0.18) | 0.026 | $8.00 \times 10^{-6}$ | 0.92 (0.79, 1.08) | 0.057 | .204 |
| IPL | -0.04 (-0.10, 0.03) | 0.026 | .173 | 0.73 (0.62, 0.85) | 0.045 | $2.30 \times 10^{-7}$ |
| HIPPO | -0.13 (-0.20, -0.06) | 0.027 | $1.00 \times 10^{-6}$ | 1.05 (0.91, 1.22) | 0.060 | .369 |
| AMYG | 0.06 (0.00, 0.13) | 0.027 | .015 | 1.01 (0.86, 1.18) | 0.061 | .892 |
| STRI | 0.36 (0.29, 0.43) | 0.027 | $2.59 \times 10^{-39}$ | 1.10 (0.97, 1.25) | 0.054 | .050 |
| THAL | -0.10 (-0.17, -0.03) | 0.027 | $2.18 \times 10^{-4}$ | 1.09 (0.94, 1.27) | 0.064 | .120 |

IDP, imaging-derived phenotype; CI, confidence interval; SE, standard error; OR, odds ratio; TCV, total cortical volume; OFC, orbitofrontal cortex; DLPFC, dorsolateral prefrontal cortex; ACC, anterior cingulate cortex; PCC, posterior cingulate cortex; INS, insula; STG, superior temporal gyrus; PREC, precuneus; IPL, inferior parietal lobule; HIPPO, hippocampus; AMYG, amygdala; STRI, striatum; THAL, thalamus.

**Table S10.** Deviations from normative values of white matter microstructural integrity (A) and their associations with aggressive behaviour (B) in individuals with schizophrenia, adjusting for current dosage of antipsychotic medication ($N$ = 549).

| | A | | | B | | |
|---|---|---|---|---|---|---|
| **IDP** | *d* (99% CI) | SE | *p* | OR (99% CI) | SE | *p* |
| GWM | -0.21 (-0.32, -0.10) | 0.043 | $7.63 \times 10^{-7}$ | 0.70 (0.57, 0.86) | 0.057 | $1.20 \times 10^{-5}$ |
| CC | -0.14 (-0.25, -0.03) | 0.043 | $9.30 \times 10^{-4}$ | 0.82 (0.65, 1.04) | 0.075 | .034 |
| CG | 0.22 (0.09, 0.036) | 0.051 | $1.80 \times 10^{-5}$ | 1.11 (0.85, 1.45) | 0.114 | .311 |
| FOF | 0.11 (0.00, 0.22) | 0.043 | .010 | 0.98 (0.74, 1.30) | 0.108 | .842 |
| SLF | 0.07 (-0.05, 0.18) | 0.043 | .128 | 1.15 (0.89, 1.47) | 0.111 | .161 |
| SS | 0.00 (-0.11, 0.11) | 0.043 | .975 | 1.08 (0.83, 1.42) | 0.114 | .438 |
| UNC | -0.10 (-0.21, 0.01) | 0.043 | .022 | 0.80 (0.61, 1.05) | 0.084 | .037 |
| CR | -0.01 (-0.12, 0.10) | 0.043 | .869 | 1.30 (0.99, 1.71) | 0.138 | .012 |
| IC | 0.27 (0.16, 0.38) | 0.043 | $6.55 \times 10^{-10}$ | 0.77 (0.64, 0.94) | 0.059 | $6.50 \times 10^{-4}$ |
| EC | 0.11 (0.00, 0.22) | 0.043 | .008 | 1.12 (0.86, 1.47) | 0.118 | .276 |
| CST | 0.04 (-0.07, 0.15) | 0.043 | .304 | 1.08 (0.84, 1.39) | 0.105 | .453 |
| PTR | -0.03 (-0.15, 0.08) | 0.043 | .417 | 0.91 (0.69, 1.20) | 0.098 | .386 |
| FX | -0.17 (-0.28, -0.05) | 0.044 | $1.46 \times 10^{-4}$ | 1.08 (0.88, 1.34) | 0.089 | .330 |
| FXST | -0.03 (-0.14, 0.08) | 0.043 | .422 | 1.01 (0.78, 1.31) | 0.101 | .928 |

IDP, imaging-derived phenotype; CI, confidence interval; SE, standard error; OR, odds ratio; GWM, global white matter; CC, corpus callosum; CG, cingulum; FOF, fronto-occipital fasciculus; SLF, superior longitudinal fasciculus; SS, sagittal stratum; UNC, uncinate fasciculus; CR, corona radiata; IC, internal capsule; EC, external capsule; CST, corticospinal tract; PTR, posterior thalamic radiations; FX, fornix; FXST, fornix stria terminalis.

**Table S11.** Deviations from normative values of grey matter volume (A) and their associations with aggressive behaviour (B) in individuals with schizophrenia, adjusting for duration of illness ($N$ = 1680).

| | A | | | B | | |
|---|---|---|---|---|---|---|
| **IDP** | $d$ (99% CI) | SE | $p$ | OR (99% CI) | SE | $p$ |
| TCV | -0.33 (-0.39, -0.27) | 0.024 | $8.89 \times 10^{-40}$ | 0.88 (0.77, 1.01) | 0.047 | .015 |
| OFC | 0.01 (-0.06, 0.07) | 0.025 | .839 | 1.08 (0.92, 1.25) | 0.064 | .217 |
| DLPFC | -0.05 (-0.11, 0.02) | 0.025 | .061 | 0.84 (0.72, 0.98) | 0.051 | .004 |
| ACC | -0.04 (-0.10, 0.02) | 0.025 | .108 | 0.96 (0.82, 1.12) | 0.057 | .470 |
| PCC | 0.08 (0.02, 0.14) | 0.024 | $9.65 \times 10^{-4}$ | 0.95 (0.82, 1.09) | 0.053 | .317 |
| INS | 0.05 (-0.01, 0.11) | 0.025 | .040 | 1.06 (0.91, 1.24) | 0.063 | .320 |
| STG | -0.08 (-0.15, -0.02) | 0.025 | .001 | 1.15 (0.99, 1.33) | 0.067 | .019 |
| PREC | 0.05 (-0.02, 0.11) | 0.025 | .061 | 1.00 (0.85, 1.17) | 0.060 | .964 |
| IPL | -0.02 (-0.08, 0.04) | 0.025 | .441 | 0.81 (0.69, 0.94) | 0.049 | $3.50 \times 10^{-4}$ |
| HIPPO | -0.07 (-0.13, 0.00) | 0.025 | .006 | 1.09 (0.94, 1.26) | 0.063 | .138 |
| AMYG | 0.09 (0.03, 0.16) | 0.025 | $1.73 \times 10^{-4}$ | 1.05 (0.90, 1.23) | 0.064 | .398 |
| STRI | 0.26 (0.11, 0.32) | 0.025 | $1.27 \times 10^{-23}$ | 1.08 (0.95, 1.22) | 0.052 | .116 |
| THAL | -0.04 (-0.10, 0.03) | 0.026 | .162 | 1.05 (0.90, 1.22) | 0.062 | .434 |

IDP, imaging-derived phenotype; CI, confidence interval; SE, standard error; OR, odds ratio; TCV, total cortical volume; OFC, orbitofrontal cortex; DLPFC, dorsolateral prefrontal cortex; ACC, anterior cingulate cortex; PCC, posterior cingulate cortex; INS, insula; STG, superior temporal gyrus; PREC, precuneus; IPL, inferior parietal lobule; HIPPO, hippocampus; AMYG, amygdala; STRI, striatum; THAL, thalamus.

**Table S12.** Deviations from normative values of white matter microstructural integrity (A) and their associations with aggressive behaviour (B) in individuals with schizophrenia, adjusting for duration of illness ($N$ = 454).

| | A | | | B | | |
|---|---|---|---|---|---|---|
| **IDP** | $d$ (99% CI) | SE | $p$ | OR (99% CI) | SE | $p$ |
| GWM | -0.13 (-0.25, 0.00) | 0.047 | .008 | 0.60 (0.47, 0.78) | 0.060 | $3.9 \times 10^{-7}$ |
| CC | 0.02 (-0.10, 0.14) | 0.047 | .633 | 0.76 (0.55, 1.04) | 0.094 | .026 |
| CG | 0.24 (0.08, 0.39) | 0.060 | $8.50 \times 10^{-5}$ | 1.08 (0.79, 1.49) | 0.133 | .518 |
| FOF | 0.17 (0.04, 0.29) | 0.047 | $4.84 \times 10^{-4}$ | 1.15 (0.79, 1.67) | 0.167 | .329 |
| SLF | 0.03 (-0.09, 0.15) | 0.047 | .505 | 1.17 (0.88, 1.57) | 0.133 | .162 |
| SS | 0.10 (-0.02, 0.22) | 0.047 | .030 | 1.10 (0.79, 1.54) | 0.144 | .460 |
| UNC | 0.01 (-0.12, 0.13) | 0.047 | .894 | 0.67 (0.46, 0.97) | 0.096 | .005 |
| CR | -0.01 (-0.13, 0.11) | 0.047 | .833 | 1.40 (0.99, 1.98) | 0.188 | .011 |
| IC | 0.20 (0.08, 0.33) | 0.047 | $1.70 \times 10^{-5}$ | 0.72 (0.57, 0.91) | 0.066 | $3.30 \times 10^{-4}$ |
| EC | 0.05 (-0.07, 0.17) | 0.047 | .307 | 1.30 (0.92, 1.85) | 0.178 | .053 |
| CST | -0.06 (-0.18, 0.06) | 0.047 | .193 | 1.19 (0.85, 1.68) | 0.158 | .181 |
| PTR | 0.01 (-0.11, 0.13) | 0.047 | .879 | 0.91 (0.63, 1.30) | 0.126 | .483 |
| FX | -0.16 (-0.28, -0.04) | 0.048 | $9.05 \times 10^{-4}$ | 1.55 (1.15, 2.08) | 0.178 | $1.50 \times 10^{-4}$ |
| FXST | 0.00 (-0.13, 0.12) | 0.047 | .938 | 0.97 (0.69, 1.35) | 0.124 | .788 |

IDP, imaging-derived phenotype; CI, confidence interval; SE, standard error; OR, odds ratio; GWM, global white matter; CC, corpus callosum; CG, cingulum; FOF, fronto-occipital fasciculus; SLF, superior longitudinal fasciculus; SS, sagittal stratum; UNC, uncinate fasciculus; CR, corona radiata; IC, internal capsule; EC, external capsule; CST, corticospinal tract; PTR, posterior thalamic radiations; FX, fornix; FXST, fornix stria terminalis.

**Table S13.** Deviations from normative values of grey matter volume (A) and their associations with aggressive behaviour (B) in individuals with schizophrenia, adjusting for lifetime substance use disorder (*N* = 778).

| | A | | | B | | |
|---|---|---|---|---|---|---|
| **IDP** | *d* (99% CI) | SE | *p* | OR (99% CI) | SE | *p* |
| TCV | -0.23 (-0.32, -0.13) | 0.036 | $5.00 \times 10^{-10}$ | 0.88 (0.75, 1.05) | 0.058 | .063 |
| OFC | 0.10 (0.00, 0.19) | 0.036 | .008 | 0.98 (0.81, 1.19) | 0.073 | .827 |
| DLPFC | 0.08 (-0.01, 0.18) | 0.036 | .019 | 0.82 (0.68, 0.99) | 0.061 | .008 |
| ACC | -0.01 (-0.10, 0.09) | 0.036 | .878 | 0.98 (0.81, 1.18) | 0.072 | .777 |
| PCC | 0.11 (0.02, 0.21) | 0.036 | .002 | 0.99 (0.82, 1.20) | 0.075 | .901 |
| INS | 0.04 (-0.05, 0.14) | 0.036 | .242 | 0.97 (0.80, 1.17) | 0.072 | .660 |
| STG | -0.15 (-0.24, -0.05) | 0.038 | $1.28 \times 10^{-4}$ | 1.03 (0.86, 1.24) | 0.073 | .665 |
| PREC | 0.16 (0.07, 0.25) | 0.036 | $8.00 \times 10^{-6}$ | 0.98 (0.81, 1.18) | 0.071 | .739 |
| IPL | 0.10 (0.01, 0.19) | 0.036 | .005 | 0.82 (0.68, 0.99) | 0.060 | .006 |
| HIPPO | -0.20 (-0.30, -0.10) | 0.038 | $1.05 \times 10^{-7}$ | 0.92 (0.76, 1.12) | 0.069 | .267 |
| AMYG | -0.06 (-0.16, 0.04) | 0.038 | .112 | 0.95 (0.77, 1.17) | 0.078 | .533 |
| STRI | 0.25 (0.15, 0.35) | 0.038 | $3.99 \times 10^{-11}$ | 1.19 (1.00, 1.42) | 0.081 | .011 |
| THAL | -0.19 (-0.29, -0.08) | 0.040 | $3.00 \times 10^{-6}$ | 0.92 (0.73, 1.15) | 0.080 | .321 |

IDP, imaging-derived phenotype; CI, confidence interval; SE, standard error; OR, odds ratio; TCV, total cortical volume; OFC, orbitofrontal cortex; DLPFC, dorsolateral prefrontal cortex; ACC, anterior cingulate cortex; PCC, posterior cingulate cortex; INS, insula; STG, superior temporal gyrus; PREC, precuneus; IPL, inferior parietal lobule; HIPPO, hippocampus; AMYG, amygdala; STRI, striatum; THAL, thalamus.

**Table S14.** Deviations from normative values of white matter microstructural integrity (A) and their associations with aggressive behaviour (B) in individuals with schizophrenia, adjusting for lifetime substance use disorder (*N* = 217).

| | A | | | B | | |
|---|---|---|---|---|---|---|
| **IDP** | *d* (99% CI) | SE | *p* | OR (99% CI) | SE | *p* |
| GWM | -0.18 (-0.36, 0.00) | 0.068 | .009 | 0.73 (0.55, 0.97) | 0.081 | .005 |
| CC | -0.15 (-0.33, 0.02) | 0.068 | .026 | 0.91 (0.70, 1.20) | 0.095 | .389 |
| CG | 0.26 (0.08, 0.43) | 0.068 | $2.04 \times 10^{-4}$ | 1.11 (0.77, 1.59) | 0.155 | .476 |
| FOF | 0.17 (0.00, 0.35) | 0.068 | .012 | 0.85 (0.59, 1.23) | 0.121 | .257 |
| SLF | 0.11 (-0.07, 0.28) | 0.068 | .121 | 1.19 (0.83, 1.72) | 0.171 | .216 |
| SS | 0.01 (-0.17, 0.18) | 0.068 | .910 | 1.13 (0.75, 1.70) | 0.179 | .429 |
| UNC | -0.11 (-0.29, 0.07) | 0.068 | .108 | 0.85 (0.63, 1.16) | 0.102 | .183 |
| CR | -0.02 (-0.20, 0.15) | 0.068 | .744 | 1.31 (0.92, 1.86) | 0.178 | .045 |
| IC | 0.40 (0.23, 0.58) | 0.068 | $1.20 \times 10^{-8}$ | 0.94 (0.67, 1.32) | 0.124 | .637 |
| EC | 0.17 (-0.01, 0.35) | 0.068 | .012 | 1.24 (0.86, 1.79) | 0.177 | .137 |
| CST | 0.08 (-0.09, 0.26) | 0.068 | .224 | 0.93 (0.65, 1.33) | 0.129 | .593 |
| PTR | -0.03 (-0.21, 0.15) | 0.068 | .664 | 0.99 (0.69, 1.41) | 0.136 | .935 |
| FX | -0.25 (-0.43, -0.08) | 0.069 | $2.52 \times 10^{-4}$ | 0.93 (0.68, 1.26) | 0.111 | .518 |
| FXST | -0.09 (-0.27, 0.09) | 0.068 | .190 | 1.12 (0.75, 1.67) | 0.174 | .472 |

IDP, imaging-derived phenotype; CI, confidence interval; SE, standard error; OR, odds ratio; GWM, global white matter; CC, corpus callosum; CG, cingulum; FOF, fronto-occipital fasciculus; SLF, superior longitudinal fasciculus; SS, sagittal stratum; UNC, uncinate fasciculus; CR, corona radiata; IC, internal capsule; EC, external capsule; CST, corticospinal tract; PTR, posterior thalamic radiations; FX, fornix; FXST, fornix stria terminalis.

**Table S15.** Deviations from normative values of grey matter volume (A) and their associations with scores on PANSS item P7 (B) in individuals with schizophrenia (*N* = 1569).

| | A | | | B | | |
|---|---|---|---|---|---|---|
| **IDP** | *d* (99% CI) | SE | *p* | OR (99% CI) | SE | *p* |
| TCV | -0.41 (-0.48, -0.34) | 0.026 | $1.33 \times 10^{-54}$ | 0.84 (0.74, 0.96) | 0.043 | .001 |
| OFC | 0.07 (0.00, 0.13) | 0.025 | .011 | 1.04 (0.90, 1.21) | 0.060 | .447 |
| DLPFC | -0.07 (-0.14, -0.01) | 0.025 | .004 | 0.83 (0.71, 0.97) | 0.049 | .002 |
| ACC | -0.06 (-0.13, 0.01) | 0.025 | .019 | 0.99 (0.85, 1.14) | 0.056 | .813 |
| PCC | 0.11 (0.04, 0.18) | 0.025 | $1.40 \times 10^{-5}$ | 0.92 (0.80, 1.06) | 0.049 | .118 |
| INS | 0.07 (0.00, 0.13) | 0.025 | .006 | 1.03 (0.89, 1.19) | 0.058 | .625 |
| STG | -0.10 (-0.17, -0.03) | 0.026 | $1.03 \times 10^{-4}$ | 1.14 (0.99, 1.32) | 0.065 | .017 |
| PREC | 0.16 (0.10, 0.23) | 0.025 | $1.20 \times 10^{-10}$ | 0.93 (0.80, 1.09) | 0.055 | .233 |
| IPL | -0.02 (-0.08, 0.05) | 0.025 | .545 | 0.75 (0.65, 0.87) | 0.044 | $7.70 \times 10^{-7}$ |
| HIPPO | -0.20 (-0.26, -0.13) | 0.026 | $7.57 \times 10^{-14}$ | 1.04 (0.90, 1.20) | 0.057 | .474 |
| AMYG | 0.03 (-0.04, 0.10) | 0.026 | .262 | 1.03 (0.88, 1.19) | 0.060 | .639 |
| STRI | 0.33 (0.26, 0.39) | 0.027 | $1.14 \times 10^{-34}$ | 1.08 (0.96, 1.23) | 0.052 | .098 |
| THAL | -0.21 (-0.28, -0.14) | 0.027 | $4.48 \times 10^{-15}$ | 1.08 (0.94, 1.26) | 0.062 | .159 |

IDP, imaging-derived phenotype; CI, confidence interval; SE, standard error; OR, odds ratio; TCV, total cortical volume; OFC, orbitofrontal cortex; DLPFC, dorsolateral prefrontal cortex; ACC, anterior cingulate cortex; PCC, posterior cingulate cortex; INS, insula; STG, superior temporal gyrus; PREC, precuneus; IPL, inferior parietal lobule; HIPPO, hippocampus; AMYG, amygdala; STRI, striatum; THAL, thalamus.

**Table S16.** Deviations from normative values of white matter microstructural integrity (A) and their associations with scores on PANSS item P7 (B) in individuals with schizophrenia (*N* = 504).

| | A | | | B | | |
|---|---|---|---|---|---|---|
| **IDP** | *d* (99% CI) | SE | *p* | OR (99% CI) | SE | *p* |
| GWM | -0.33 (-0.44, -0.21) | 0.046 | $9.99 \times 10^{-13}$ | 0.77 (0.62, 0.94) | 0.061 | .001 |
| CC | -0.18 (-0.29, -0.06) | 0.045 | $8.20 \times 10^{-5}$ | 0.83 (0.66, 1.05) | 0.076 | .047 |
| CG | 0.33 (0.19, 0.48) | 0.056 | $2.79 \times 10^{-9}$ | 1.14 (0.86, 1.52) | 0.125 | .217 |
| FOF | 0.16 (0.04, 0.27) | 0.045 | $5.84 \times 10^{-4}$ | 0.91 (0.67, 1.23) | 0.107 | .412 |
| SLF | 0.12 (0.01, 0.24) | 0.045 | .005 | 1.11 (0.87, 1.40) | 0.102 | .277 |
| SS | 0.04 (-0.07, 0.16) | 0.045 | .348 | 1.05 (0.80, 1.36) | 0.107 | .663 |
| UNC | -0.06 (-0.18, 0.05) | 0.045 | .147 | 0.81 (0.62, 1.05) | 0.084 | .039 |
| CR | 0.04 (-0.07, 0.16) | 0.045 | .357 | 1.26 (0.96, 1.65) | 0.133 | .028 |
| IC | 0.34 (0.22, 0.46) | 0.046 | $1.31 \times 10^{-13}$ | 0.73 (0.60, 0.89) | 0.055 | $3.40 \times 10^{-5}$ |
| EC | 0.16 (0.04, 0.27) | 0.045 | $3.95 \times 10^{-4}$ | 1.03 (0.78, 1.35) | 0.108 | .797 |
| CST | 0.06 (-0.05, 0.18) | 0.045 | .171 | 1.12 (0.87, 1.45) | 0.111 | .251 |
| PTR | 0.00 (-0.12, 0.11) | 0.045 | .954 | 0.89 (0.67, 1.17) | 0.096 | .265 |
| FX | -0.24 (-0.36, -0.12) | 0.046 | $1.36 \times 10^{-7}$ | 1.13 (0.91, 1.39) | 0.092 | .147 |
| FXST | -0.03 (-0.15, 0.08) | 0.045 | .482 | 1.01 (0.78, 1.30) | 0.100 | .941 |

IDP, imaging-derived phenotype; CI, confidence interval; SE, standard error; OR, odds ratio; GWM, global white matter; CC, corpus callosum; CG, cingulum; FOF, fronto-occipital fasciculus; SLF, superior longitudinal fasciculus; SS, sagittal stratum; UNC, uncinate fasciculus; CR, corona radiata; IC, internal capsule; EC, external capsule; CST, corticospinal tract; PTR, posterior thalamic radiations; FX, fornix; FXST, fornix stria terminalis.
